# Hump nosed pit viper envenoming in Coastal Karnataka- unravelling the centuries of deadly camouflage

**DOI:** 10.64898/2026.03.05.26347697

**Authors:** Freston Marc Sirur, Vrinda Lath, Devika Jabagodu Lingappa, Ramya, Nirmal U Kulkarni, Asha Kamath, Usha Wagle

**Affiliations:** Department of Emergency Medicine, Kasturba Medical College, Manipal Academy of Higher Education, Manipal, India; Department of Emergency Medical Technology, Manipal College of Health Professions, Manipal Academy of Higher Education, Manipal, India; EMS Instructor, Red Tact Academy for clinical training, Bangalore, Karnataka, India; Department of Emergency and Trauma Care Technology, BGS Global Institute of Allied Health Science, Kengeri, Bangalore, Karnataka, India; Mhadei Research Center, Viper Specialist Group, IUCN 6, Hiru Naik Bldg, Dhuler, Mapusa, Goa- 403507, India; Department of Applied Statistics & Data Science, Prasanna School of Public Health, Manipal Academy of Higher Education, Manipal, India

**Keywords:** Hump nosed pit viper, snakebite, antivenom, *Hypnale-hypnale*, Venom

## Abstract

**Background:** The Hump-nosed pit viper is a recognized but neglected medically significant species causing morbidity and mortality, with non-availability of a specific antivenom. There are many gaps in our understanding of its envenomation, including burden, clinical syndrome, complications and management.

**Methodology:** The study is a retrospective sub analysis of the Prospective VENOMS registry and hospital records of Hump Nosed Pit Viper envenomation from a single tertiary care center in coastal Karnataka from May 2018 to March 2024. Epidemiology, syndrome, complications and treatment strategies have been described. A linear mixed model analysis was conducted to study the effect of different therapeutic interventions in combating venom induced consumptive coagulopathy (VICC)

**Principal Findings:** Of 46 cases, 24 patients had VICC. The most common complications were AKI (21.7%), TMA (10.9%) and stroke (4.4%). Anaphylaxis to ASV (23.9%) was the most common therapeutic complication. Therapeutic interventions included ASV, administration of blood products and therapeutic plasma exchange along with supportive care. The linear mixed model revealed that administration of blood products (p=<0.001) had the strongest influence on the INR value, however, often resulting in a transient decline in INR value. ASV (p=0.052) caused only marginally significant change in INR. The role of TPE could not be statistically inferred, however, individual cases with severe VICC improved without complications, therefore it required further study but can be considered in critical cases.

**Conclusions/Significance:** This study describes the syndrome of hump-nosed pit viper envenomation, while highlighting the urgent need for a species-specific antivenom, recommends treatment strategies that can be used in the interim. Additionally, geo-spatial mapping draws attention to hotspots and the hypothesis that HNPV in coastal Karnataka have regionally distinct toxicity trends.

**Author Summary:** India is often known as the snakebite capital of the world, with recent literature suggesting that not all death and disability is attributable to the “Big four” and highlighting regionally significant species of snake. In the Western Ghat region of India, envenomation by the Hump-nosed pit viper is increasingly being reported. In coastal Karnataka, it has been reported as the second most common cause of envenomation following the Russell’s Viper, causing systemic envenomation and death. However, little is known about why envenomation is common in this region, is it increasing, how to diagnose envenomation, its clinical syndrome, the anticipated complications, and most importantly, an effective treatment strategy. This study reports envenomation in 46 patients, resulting in 3 deaths, and 24 patients developing derangement in coagulation parameters.

## Introduction

The Hump-nosed pit viper, only recently recognized in medical literature as a medically significant species, is endemic to the Western Ghats of India and Sri Lanka.^1–5^. Until 2018 the hump nosed pit viper (HNPV) was underrecognized as a medically significant species in the state of Karnataka with no cases of envenoming published in scientific literature in the state. A few reports recognized its presence, and that envenoming could take place^6,7^. One of the primary reasons is misidentification or no identification of the culprit species by victims, the public and treating physicians. This problem has been compounded by the absence of a reliable diagnostic kit. The first confirmed cases from Karnataka published in scientific literature in 2022 reported all three cases as critically ill with two deaths and inefficacy of available polyvalent anti-venom^1^. Since 2019 the VENOMS Registry has been recording cases in an organized format with a focus on identifying gaps and best practices in pre-hospital, in-hospital, diagnostics including snake species identification, treatment protocols and knowledge of public on snake envenoming. A strong focus on HNPV has been given over the years and this study reflects 5 years of data collected just before the start of the registry up until March 2024. India has since long been labelled as the snakebite capital of the World with the “Big Four” taking center stage. However, in recent times regional expertise and focused studies have identified regionally significant venomous species, new venomous species, and importantly the demonstration of clinical ineffectiveness of anti-venom. It must be presumed that many of these species have existed for millennia but somehow escaped the eyes of medical science for numerous reasons. This study deepens our understanding of Hump-nosed pit viper envenoming in coastal Karnataka. It sheds light on the syndrome, morbidity and mortality and treatment strategies. The One Health perspective helps us to scope viable solutions at the grassroot and through the registry, align implementation of solutions with the objectives of the National Action Plan for Snakebite Envenoming^8^.

## 2. Methods

### 2.1 Study design

The study is a retrospective analysis of Hump Nosed Pit Viper envenomation recorded in a prospective registry from a single tertiary care center in coastal Karnataka from May 2018 to March 2024. Data was collected by screening hospital records, and from the VENOMS Registry. Cases prior to the VENOMS registry were collected by screening hospital records, and cases with culprit species recorded as hump nosed pit viper were considered for analysis.

From the VENOMS Registry epidemiological, clinical, pre-hospital, complications, and treatment related data were sourced. The method of identification described below was followed, and photographic records of culprit specimens are maintained.

Methods of identification: The following methods of identification were used for this study, in descending order of quality of evidence.

a. Killed or live specimen produced at the hospital In cases where a live or dead specimen was brought to the hospital, scale counts were not performed due to safety concerns and lack of expertise in scale count assessment. Instead, clear photographs of the specimen were obtained and later verified by experts for species identification. All live specimens brought to the hospital were safely released back into their natural habitat in coordination with the Forest Department.
b. Photographic/videographic evidence captured of the culprit snake Cases with evidence, either photograph, killed or live specimen, were identified based on morphological characteristics by the authors (FMS, VL), who have expertise through training during herpetological expeditions (Pit Viper Expeditions by Herpactive, Centre for Wilderness Medicine Expeditions). These were also verified by NK. Scale counts could not be performed due to the damaged condition of specimens or the quality of the photographic / videographic evidence.
c. Syndromic identification In Hump-nosed pit viper envenomation, clinical syndrome in the presence of coagulopathy combined with geographical and habitat information, is a reliable method of identification in our region when response to Indian Polyvalent ASV is not observed^1,5^.
d. Photographic identification has not been considered in this series as the HNPV is often mistaken for other viperine species from this region, particularly the Russell’s viper and brown morph of the Malabar pit Viper.

### 2.2 Ethics statement

The study was approved by the Kasturba Hospital Institutional Ethic Committee (IEC1:34/2024). The VENOMS registry is a prospective, Clinical Trials Registry – India (CTRI-https://ctri.nic.in/Clinicaltrials/login.php) registered, multi-centric hospital-based registry on all envenomation (CTRI/2019/10/021828). All patients enrolled in the registry provided informed consent for participation. Cases included prior to initiation of registry were included after obtaining necessary permissions. Written informed consent was obtained from the patient or a legally authorized representative, for collection of data including specimens and bite site images. For pediatric patients, written consent was obtained from parents or legal guardians.

### 2.3 Treatment protocol

As there is no specific treatment protocol for HNPV envenomation, supportive treatment was administered as per the standard treatment recommended by the National Standard Treatment Guidelines^9^ and the World Health Organization South-East Asia Region (WHO SEARO) guidelines^10^. Indian Polyvalent ASV when administered, was either prior to being referred to our center or before identity of the culprit species was confirmed. As an emergency department protocol, ASV is withheld once evidence of the culprit species was confirmed either by specimen or photographic evidence. If evidence is not available, identification of HNPV is challenging, unless the patient develops VICC that does not improve with administration of ASV in this region.

### 2.4 Statistical methods

Descriptive statistics were used, with qualitative data represented as percentage and quantitative data represented as mean and standard deviation. To analyze the effects of different interventions on the INR value, linear mixed model analysis was performed. P-values were calculated to assess the likelihood that the observed differences occurred randomly, with a threshold of p < 0.05 indicating a less than 5% probability of such an occurrence by chance. Parameters with p-values below this threshold were deemed statistically significant.

Analytical tools- The data was captured as per the standard proforma of the VENOMS registry, tabulated and analyzed on Microsoft excel (version 2410) and Jamovi (version 2.7.5). Graphs were generated on Jamovi (version 2.7.5) and Python (version 3.12.3). GIS mapping was done using Qgis and the state outline map was taken from Karnataka Geographic Information System.

## 3. Results

All 46 cases are within the recorded habitat of the HNPV (Fig 1). Most of the non-critical cases appear to be clustered around the study center marked red, whereas two cases with fatality and one moribund patient marked in black were from the northern part of the Udupi district and North Karnataka (Uttara Kannada) district.

**Fig 1:**
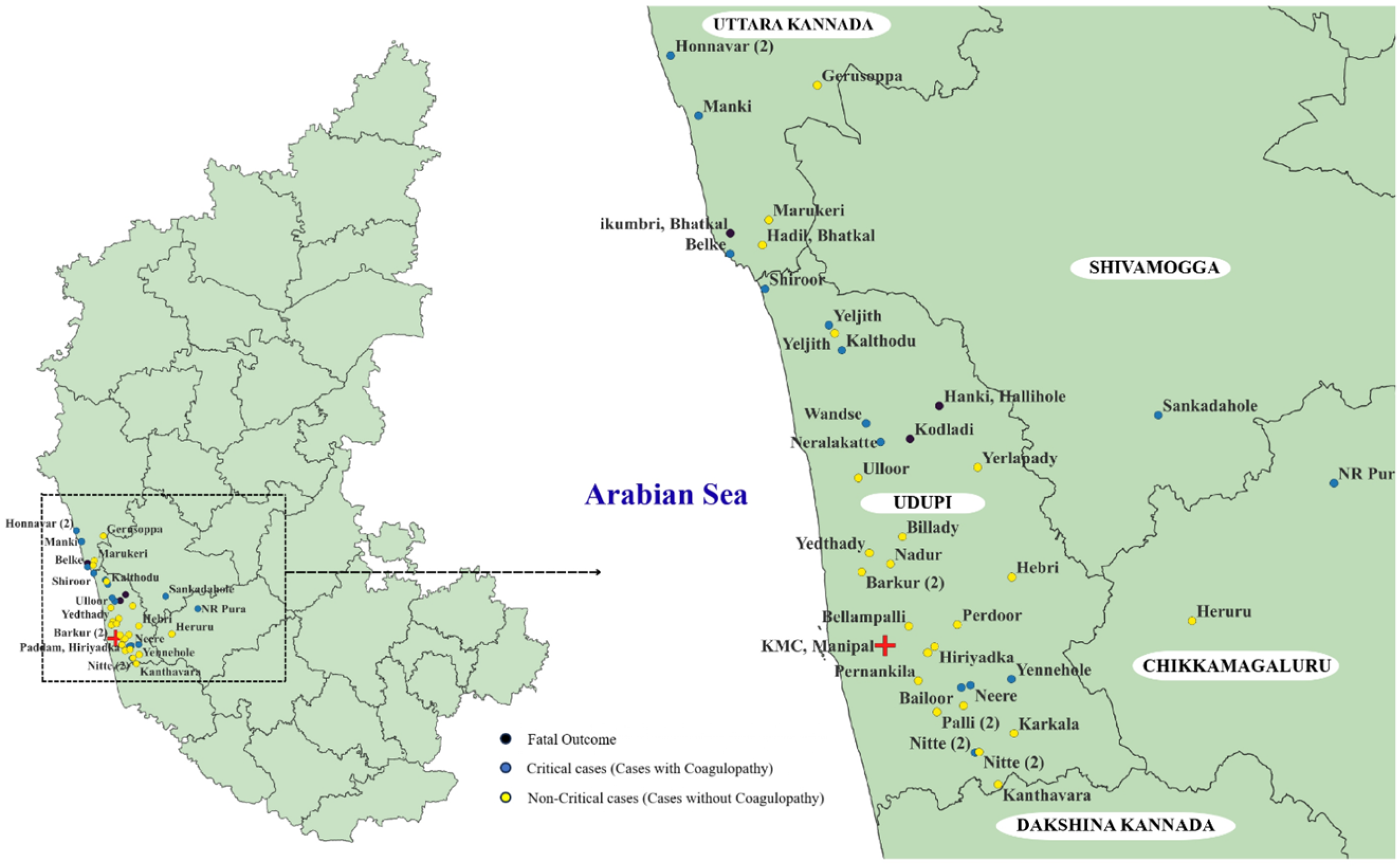
Geospatial mapping. The base shapefile used for district boundaries was sourced from the GitHub repository (District.zip, available at: https://github.com/JKAY3366/karnataka_pop_by_hr), which is licensed under CC BY 4.0. All mapping and spatial analyses were performed using QGIS software.

Fig 2 depicts images of different morphs of some culprit specimens from different parts of Karnataka.

**Fig 2:**
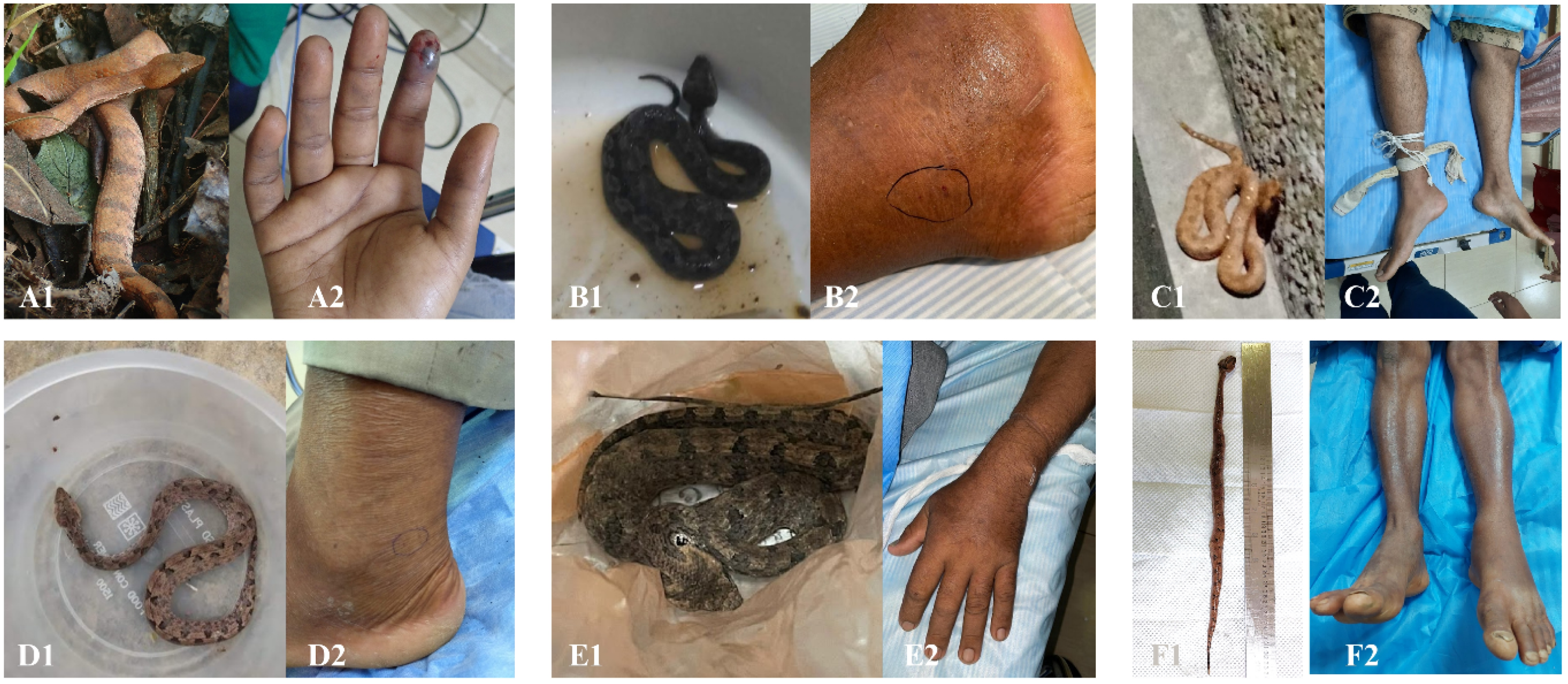
Images of culprit specimens with bite sites.

As shown in the Table 1, of the 46 cases of Hump-nosed pit viper envenomation, 26 (56.5%) were male. While most patients were between the ages of 30-70 years (78%), 10.9 percent of the cases were children aged <10 years old. Bites occur throughout the year, with a majority during summer and post-monsoon/winter. All patients had bites over the extremities, either sustained while walking where they accidentally stepped on the snake, or when picking up leaves or twigs for clearing, firewood or fodder. While agricultural activity accounted for the highest number of cases, many patients were bitten during routine activities outside their residence, collecting firewood, playing, and cleaning their yards. Most cases (52.1%) arrived at our tertiary care center within 6 hours of the bite. Most cases were referred to our center from peripheral centers

**Table 1:**
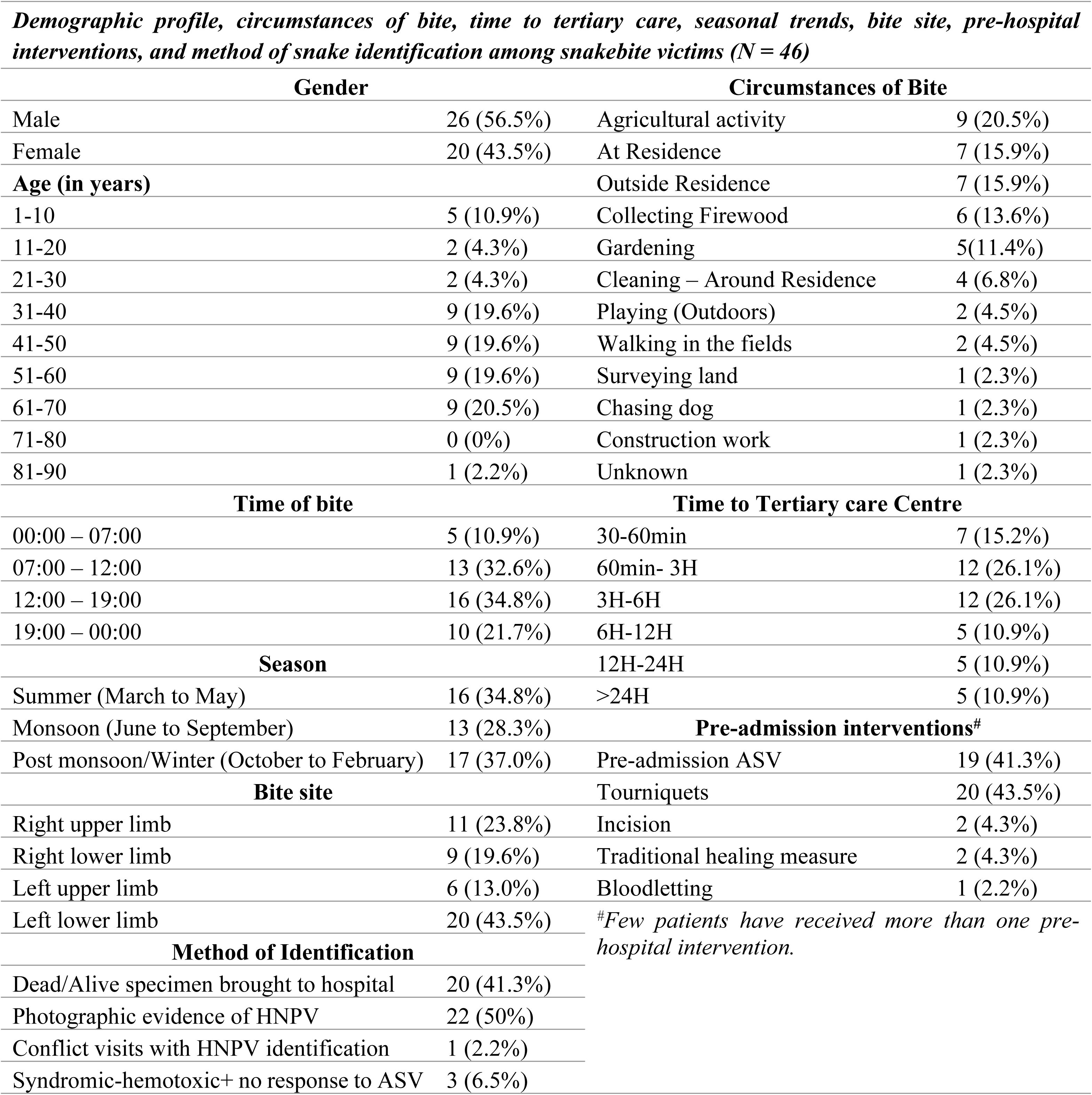
Demographic profile, circumstances of bite, time to tertiary care, seasonal trends, bite site, pre-hospital interventions, and method of snake identification.

43 cases presented with evidence in the form of dead or live specimens brought to the hospital, or a photograph of the culprit specimen. One case required a site visit to rescue and confirm the identification of the culprit species at the victim’s residence. Tourniquet was applied in 43.5% of cases. Only 2 patients visited traditional healers. In 41.3% of cases, ASV was administered at peripheral centers prior to transfer to our center. Doses varied from a test dose to a minimum of 4 and a maximum of 40 vials.

As shown in Table 2, of the 46 cases, 19 had only local envenomation in the form of pain and swelling at the bite site with some experiencing bleeding and discoloration of the bitten limb. 24 patients had both local signs in the form of pain, swelling at bite site and systemic envenomation in the form of coagulopathy, evidenced either by derangement in INR value or by TEG. One patient had a normal INR and hypo coagulable TEG. 3 patients did not have any signs of envenomation. Coagulation classification was attempted for 24 cases who has coagulopathy. The ranges used for PT classification were derived from the hematologic component of the Snakebite Severity Score (SSS) developed and validated by Dart et al. In our cohort, 33.3% of patients had PT <20 seconds, 29.2% had PT between 20–50 seconds, and 37.5% had PT >120 seconds. We did not apply the full SSS to score patients, as this system was specifically designed for crotalid (pit viper) envenomation and may not be directly applicable to our study population. The most common complication was AKI (21.7%) followed by Thrombotic Microangiopathy (TMA) (10.9%) characterized by thrombocytopenia and the presence of schistocytes^11^. Two patients developed stroke- one ischemic and one hemorrhagic. Anaphylaxis to ASV was seen in 11 cases (23.9%) of which five (11.4%) occurred at peripheral centers. Six (13%) patients underwent 1 to 4 cycles of therapeutic plasma exchange (TPE) and six (13%) required hemodialysis, with the number of sessions ranging from 1 to 12. Among the 10 patients who developed AKI, eight had associated VICC, while two had no evidence of coagulopathy. Only two patients required surgical intervention for local complications, one being a pediatric patient.

**Table 2:** Clinical presentations, complications, therapeutic interventions, patient outcomes, and hospitalization duration among snakebite victims.

| **Clinical presentation** |  | **Therapeutic Interventions\*** |  |
| --- | --- | --- | --- |
| Local signs of envenomation only | 19 (41.3%) | Endotracheal intubation | 4 (8.7%) |
| Local & Systemic signs of envenomation | 24 (52.2%) | Inotropic support | 1 (2.2%) |
| No signs of envenomation | 3 (6.5%) | ASV | 30 (65.2%) |
| Shock on arrival | 2 (4.35%) | Blood Products | 15 (32.6%) |
| **Coagulation classification** |  | Therapeutic plasma exchange | 6 (13%) |
| • PT<20seconds | 8 (33.3%) | Hemodialysis | 6 (13%) |
| • PT<20sec to 50 sec | 7 (29.2%) | Fasciotomy | 1 (2.2%) |
| • PT>120sec | 9 (37.5%) | Debridement | 1 (2.2%) |
| **Complications** |  | **Patient outcome** |  |
| Pre-admission anaphylaxis | 5 (11.4%) | Discharge with full recovery | 35 (76.1%) |
| In-hospital Anaphylaxis | 6 (13.63%) | Discharge with Disability | 4 (8.7%) |
| Acute Kidney injury (HD) | 10 (21.7%) | Death | 2 (4.3%) |
| Ischemic stroke | 1 (2.2%) | DAMA | 5 (10.9%) |
| Intracranial Bleed | 1 (2.2%) | **Duration of hospitalization** |  |
| TMA | 5 (10.9%) | Mean | 6.5 |
| Compartment syndrome | 1 (2.2%) | Maximum days | 28 |
| Acute coronary syndrome | 1 (2.2%) | Minimum days | 1 |
| Acute Pulmonary oedema | 2 (4.3%) |  |  |
| Encephalopathy | 1 (2.2%) |  |  |
| **AKI Cases Stratified by Coagulation Profile** |  | *PT-Prothrombin Time* |  |
| AKI + PT<20sec | 4 (40%) | *TMA- Thrombotic microangiopathy* |  |
| AKI + PT<20sec to 50 sec | 2 (20%) | *ASV – Anti-snake Venom* |  |
| AKI + PT>120sec | 4 (40%) | *DAMA- Discharge Against Medical Advice* |  |

While 76.1% of the patients recovered completely, 4 (8.7%) were discharged with disability, which included persistent renal failure and local complications such as non-healing ulcers and surgical scars. Two patients passed away at our center while one moribund patient with an ischemic stroke passed away at a local center. Complications resulting in death were strokes- ischemic and hemorrhagic, and cardiac failure with acute kidney injury.

23 patients had abnormal INR values which are plotted against time (Fig 3). 13 (28.26%) patients had INR values above 4, of which 9 (19.56%) were >11. Notably, INR values remained high even after 6 days following envenomation in some cases

**Fig 3:**
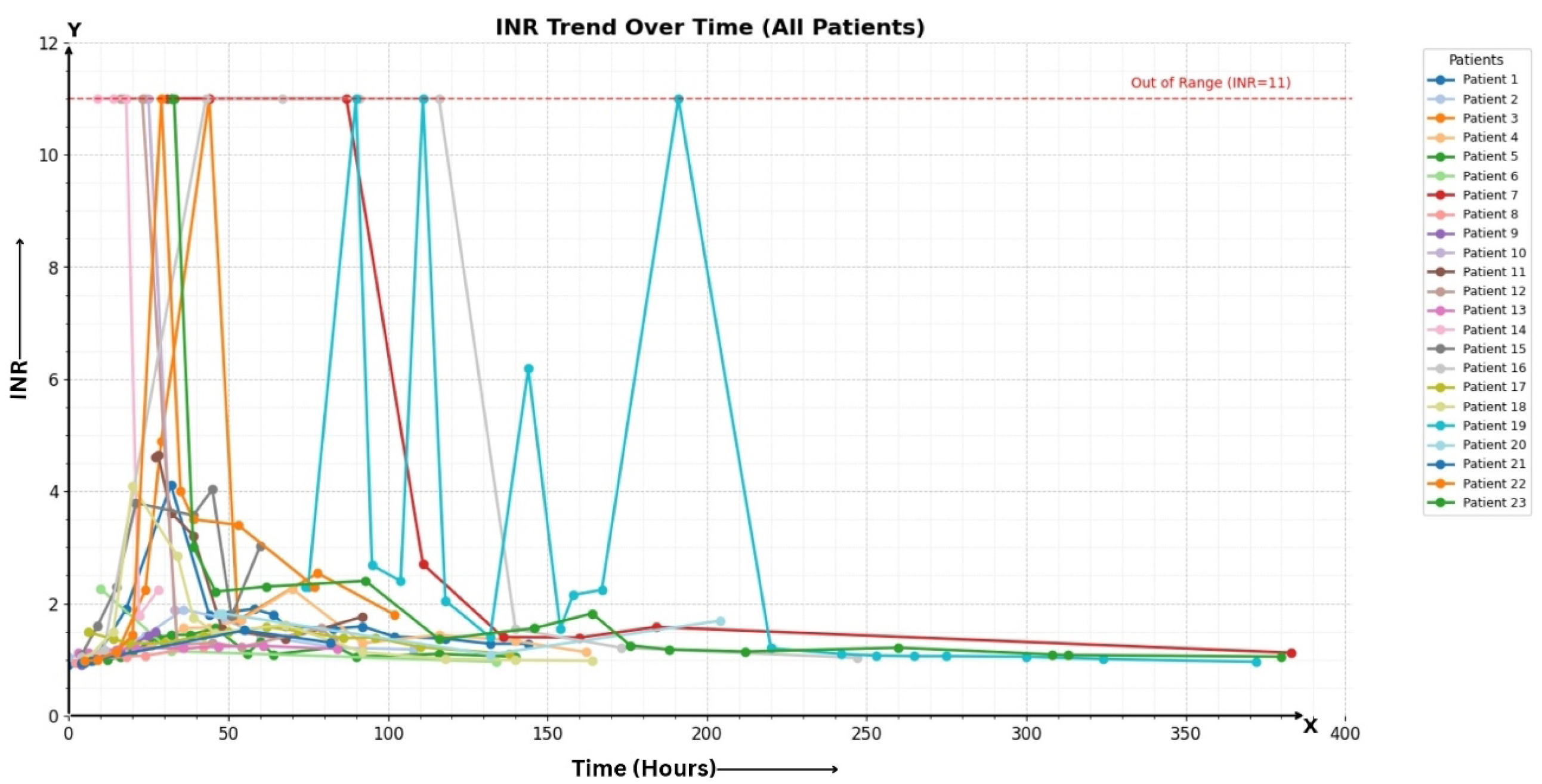
Line plot of INR vs Time in cases with coagulopathy.

As shown in Table 3, of the 23 cases with derangement in INR, 22 cases were included in the linear mixed model analysis, where the effects of interventions- Administration of ASV, Blood products and Therapeutic Plasma Exchange (TPE) on the INR values were studied. The Log INR value was considered as a dependent variable, blood products and TPE as factors, time and vials of ASV as covariates and patient ID as cluster variables. The patient ID is considered as the random effect, while blood products, ASV, TPE, and Time are considered as fixed effects.

**Table 3:** Linear mixed model results comparing effects of ASV, TPE, Blood products and Time on log INR value.

| Parameter Estimates (Fixed coefficients) |  |  |  |  |  |  |  |  |
| --- | --- | --- | --- | --- | --- | --- | --- | --- |
| 95% Confidence Intervals |  |  |  |  |  |  |  |  |
| Names | Effect | Estimate | SE | Lower | Upper | df | t | p |
| (Intercept) | (Intercept) | 0.83705 | 0.13595 | 0.56889 | 1.10522 | 59.6 | 6.157 | <.001 |
| Blood products | 1 - 0 | 0.55368 | 0.11424 | 0.32834 | 0.77903 | 191.4 | 4.847 | <.001 |
| TPE | 1 - 0 | 0.08691 | 0.19498 | -0.29768 | 0.47151 | 186.1 | 0.446 | 0.656 |
| Time | Time (INR) | -0.00258 | 7.02e-4 | -0.00396 | -0.00119 | 180.5 | -3.671 | <.001 |
| ASV | ASV | 0.01388 | 0.00710 | -1.34e-4 | 0.02788 | 184.6 | 1.954 | 0.052 |

The marginal R^2^ was found to be 0.190 (p< 0.001) and conditional R^2^ was 0.462 (p< 0.001) indicating that 46.2% of the total variance in log INR was accounted for by both fixed effects (time, ASV, blood products and TPE) and random effects (individual patient differences), while the fixed effects accounted for 19% of total variance of log INR. Both conditional and marginal effects had a high LRT X^2^, 71.8 and 52.37 (p<0.001). An Inter Class Correlation (ICC) of 0.336 indicates that 33.6% of the variances in log INR is attributable to the effect of individual patients’ underlying characteristics on the variance in log INR.

Based on this, Time (Estimate = −0.0025) and administration of blood products (Estimate = 0.55) are statistically significant variables that change the log INR values (p<0.001) whereas TPE(p=0.656) did not cause a statistically significant change and ASV (p = 0.052) caused a marginally significant change.

When plotting Log INR against time for each intervention i.e., TPE, Blood products an ASV (Fig 4a, 4b, and 4c), it is noted that in the patients who underwent TPE (n= 6) INR follows a similar trend to that over time (Figure 4a) whereas, those who received blood products saw a decrease in INR with vertical jumps, indicating that the INR values transiently decrease with blood products. In the patients who were administered ASV, the decline in INR is similar to that over time.

**Figure 4:**
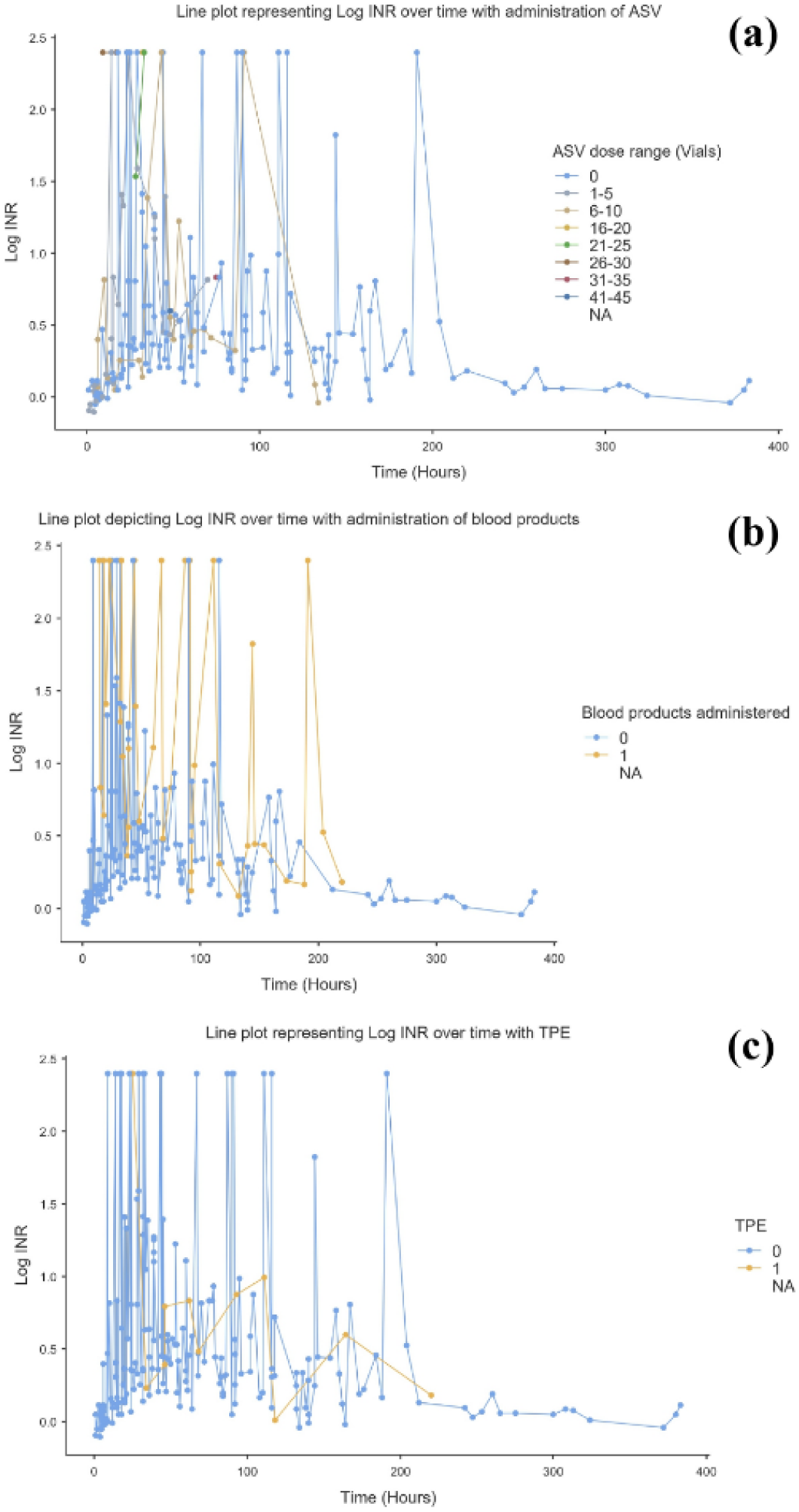
Graphical representation of effects of different interventions on INR over time. (a) Line plots representing Log INR over time with administration of ASV (b) Line plots representing Log INR over time with administration of blood products (c) Line plots representing Log INR over time with TPE

Figure 5 illustrates the temporal trends in INR in relation to administration of antivenom (ASV), therapeutic plasma exchange (TPE), and blood products in patients with severe envenomation–associated VICC. A total of six patients underwent therapeutic plasma exchange; representative INR trends of three patients are depicted in the figure. Despite initial ASV administration, INR remained markedly elevated in all cases. Following initiation of TPE, a consistent and rapid decline in INR was observed, with progressive normalization over subsequent hours. This trend was seen across all illustrated cases, even when blood products were used adjunctively. These findings demonstrate that timely initiation of TPE is associated with accelerated resolution of coagulopathy in severe VICC.

**Fig 5:**
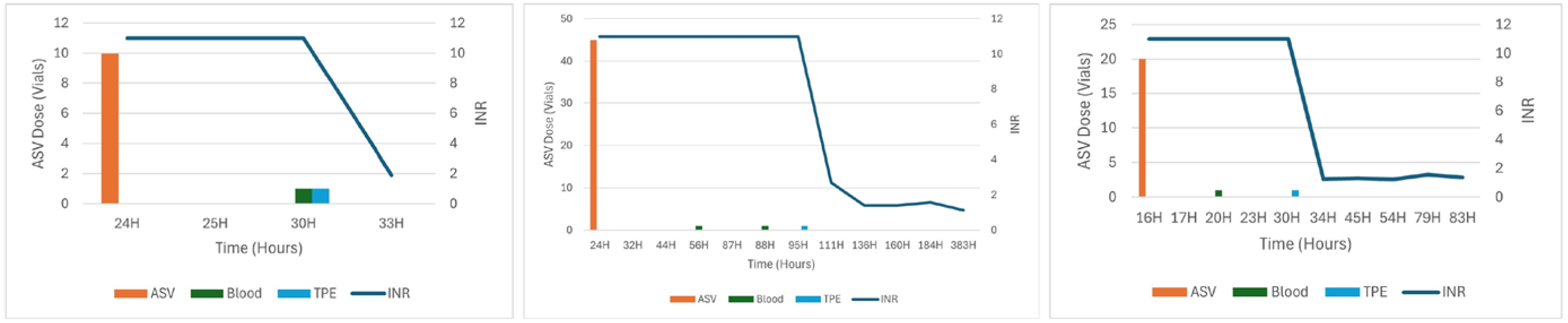
Trends in INR with ASV, Plasma exchange and Blood products.

## 4. Discussion

*Species description and range-* The hump nosed pit viper ranges from Sri Lanka into the Indian mainland where its range extends from the Western Ghats of Tamil Nadu and Kerala into Karnataka, Goa and southern reaches of the Western Ghats of Maharashtra. Three species (Hypnale *hypnale, hypnale zara and hypnale nepa)* are described all of which are found in Sri Lanka and only one, (*Hypnale hypnale*) is found in India (Fig 6)

**Fig 6:**
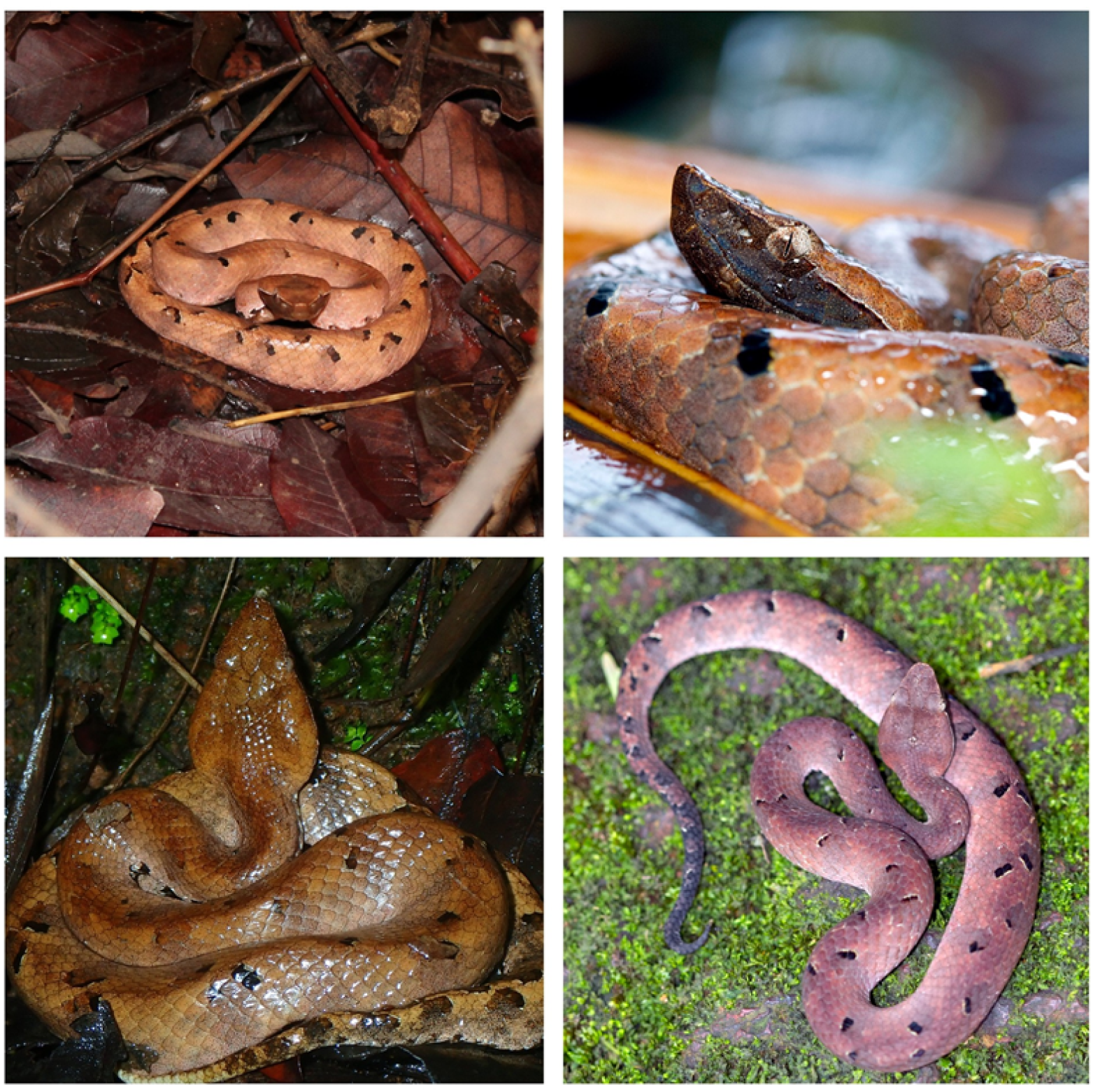
Representative images of Hump-nosed pit vipers.

In India, there are three color variations known-

a. Deep orange above with V markings interspersed with large angular or oblong spots- adults
b. Brownish or dark brownish with intricate V markings and whitish tail- juveniles
c. Grayish brown above with lateral chocolate brown spots that fade with age- adults

All these variations have pale brown underside with speckled brown or heavily powdered brown spots depending on the age (more spots on younger individuals).

*Habit, habitat and Interactions -* While limited to the Western Ghat region, their preferred habitat ranges from human habitation (mixed areca, cardamom, rubber and banana plantations) to mixed moist deciduous tropical wet evergreen and semi-evergreen forests with a marked preference for canebrakes in Goa and Karnataka.^12^ It has been sighted in disturbed habitats and secondary vegetation too above ground, besides being found in leaf litter and near water bodies perched on rotting logs. Its preferred prey is geckos, skinks and smaller rodents, all of which are plenty on the forest floor. Viviparous in nature, Hypnale gives birth to 4-18 young ones, and these possess venom at birth. Masters in camouflage, the young ones lay in coils and lure their prey with the help of a whitish tail tip that they vibrate, thus mimicking a worm that attracts geckos and skinks. Once in striking distance the strike is quick and fatal. Young ones are a dark orange color with clear coffee brown inverted V markings and are known to strike with the least amount of provocation, causing more bites in plantations.

Locally common in areas where it is found, densities have not been studied in detail. However, the number of individual sightings depends on habitats and rainfall patterns. Monsoon surveys have recorded individuals on roads, trekking paths, and in stored firewood.

As noted in our study, many bites occurred outside of agricultural fields, in habitats likely in the forests or fringes where they lie camouflaged. While mostly nocturnal, they are often seen during dawn and dusk. In our study, many bites occurred in the evening or at night, also perhaps coinciding with human activity.

Bite cases appear to have increased over the years. This may be explained either by increased awareness and better reporting in the region, or increased pressure on their habitat, resulting in more human activity such as forestry, plantations and the building of homes and farms. However, there is still a stark under-reporting, which can be described somewhat akin to an *Iceberg Phenomenon.* Not all healthcare providers can reliably identify the species. Due to its habit, it is often not seen, with patients recognizing the bite much later. Even if seen, it shares habitat with three other similar looking viperine species in this region- juvenile Russell’s viper, Saw-scaled viper and brown morph of the Malabar pit viper, which results in misidentification. Patients who present without evidence of culprit species- classified based on syndrome as viperine envenomation, are treated as per standard of care with ASV. The identification of HNPV is only possible in patients with severe coagulation derangement which does not show any improvement with ASV. Many patients only develop local envenomation and may therefore be treated at peripheral centers or may not seek healthcare altogether. A lack of reliable point-of-care diagnostic tools contributes to the underestimation of its burden. In our region, the reported incidence is second only to Russell’s viper envenomation^13^. It is important to note that Russell’s viper is dynamic and can be found across various habitats whereas the others are habitat specific. The knowledge of their natural behavior and habitat overlap are key elements in determining which species is likely to have bitten a patient even before the onset of signs and symptoms in some cases. This knowledge base is important for all healthcare workers in the Western Ghats for numerous reasons including the treatment of patients, developing targeted anti-venoms, human-wildlife conflict mitigation, prevention, effects of climate change on habitats and behaviors, and risk stratification.

### Venomics and Antivenomics

Proteomic studies have been performed on the Hypnale species from Sri Lanka and India. The studies on specimens from Sri Lanka revealed an abundance of SVMP (36.9%) LAAO (11.9%) and PLA2 (40.1%) ^14^, however, the venom proteome from the Karnataka Western Ghats revealed SVSP (44.4%) and CRISP (23.1%) as the more abundant components as opposed to the relatively lower abundance of PLA2 and SVMP (1-5%)^15^. Based on the venom composition, SVSPs act as a catalyst in the coagulation cascade, contributing to consumptive coagulopathy and hemorrhagic symptoms, while CRISP results in pro-inflammatory state and affects wound healing. In-vitro studies corroborate clinical observations of poor efficacy of the Indian polyvalent ASV against the “Big Four” in envenomation by Hump nosed pit viper. While the pentavalent antivenom from Sri Lanka which covers Hypnale species, has demonstrated better binding in vitro, it is yet to be tested in India in vivo^15,16^. Antivenom against *C rhodostoma* has been shown to effect cross neutralization, but this too is only experimental^17^. While monovalent antivenom has been developed, it has only been tested in vitro^18,19^. It may be anticipated that its effectiveness may differ considering the different venom profiles in both regions.

### Clinical Features

Our study reports a higher incidence of coagulopathy compared to studies from Sri Lanka as well as Kerala^2,3,5,20^.

### Morbidity and mortality

- As per our study, the case fatality rate is 4.3%. One patient passed away at a local hospital, bringing the total deaths to 3 (6.5%). Sajeeth Kumar et al in Kerala recorded 1.3% mortality^21^, Siju V Abraham et al reported 1 (2.4%)^5^. while in Sri Lanka deaths were reported in 1.3-1.7% of cases^20,22,23^. In Karnataka our study is the first one reporting HNPV deaths, while in Goa no reports exist. However, FMS has been involved in telemedical support for at least 3 cases on HNPV envenoming. However, verbal informal surveys did not indicate any recorded deaths or severe envenoming from HNPV in Goa so far. Cases have been reported from the Western Ghat region of Tamil Nadu but larger studies across its range will be needed to truly quantify death due to HNPV envenomation. Anaphylaxis to ASV was seen in (23.9%) of which 5 (11.4%) occurred at peripheral centers. In case of confirmed HNPV envenomation, this is an unacceptable risk, particularly as it may result in airway compromise and shock in the pre-hospital and primary care setting where advanced resuscitation capabilities are limited. Clearly, the risks outweigh the benefits in administering ASV. 21.7% of Acute Kidney Injury with 13% of patients needing Emergency Dialysis and 13% of patients undergoing Therapeutic plasma Exchange. While AKI and TMA complicated a larger number of cases, the complications of stroke-ischemic and hemorrhagic, as well as cardiac (2.2%)- likely myocardial infarction or myocarditis with pulmonary edema and AKI resulted in death.
- Reported complication of HNPV from India and Sri Lanka include Stroke, myocarditis, acute Kidney injury, Micro angiopathic hemolytic anemia, systemic bleeding, hyponatremia, seizures, hemolytic uremic syndrome, thrombotic thrombocytopenic purpura, pulmonary hemorrhage, Kounis syndrome and purpura fulmians^1,2,4,20,22–25^.
- Of the 93.5% of patients that developed signs of local envenomation, most were managed conservatively, with only 2 patients requiring surgical intervention in the form of debridement and fasciotomy.

### Treatment strategies- current practices

Polyvalent ASV manufactured by Bharath Serums did not demonstrate clinical effectiveness for all cases with severe VICC, local effects, mild to moderate envenoming or subclinical coagulopathy. While it is not recommended or mandated by protocol to administer available ASV in HNPV envenoming, many physicians who are not yet fully acquainted with it are still administering ASV claiming para specificity, or due to lack of confirmation of identity of culprit species. While standard guidelines do not mention a specific treatment protocol, administration of ASV is not recommended and only supportive care is recommended. Studies have suggested a benefit of therapeutic plasma exchange^26^. Blood products are administered in cases of severe coagulation derangement. While there is a concern that they may worsen consumptive coagulopathy, studies have demonstrated a more rapid correction of coagulopathy, but no clear benefit^27,28^. Added risks of TRALI and TACO need to be considered.

An ideal treatment protocol is still debatable, with mounting evidence against ASV. We performed a linear mixed model analysis to study the effect of each intervention on the INR in the patients with coagulopathy.

The choice of using a linear mixed model for studying the effects of different interventions on the value of INR is justified by the model fit and random component analysis. While time, ASV, blood products and TPE individually affect the INR values, individual patient factors appear to account for a large variance in the log INR values. Based on the results of the model, administration of blood products (Estimate = 0.55) and time (Estimate = −0.0025) significantly influenced the INR values (p=<0.001). The decline of INR values over time indicates the natural resolution of VICC in the body. However, prolonged effects of venom can result in complications, including renal injury, TMA and even stroke. Therefore, we study the effects of other interventions. The administration of all interventions at very high INR values suggests that the interventions were used in the most severe cases of VICC. However, this may be a confounding factor in the interpretation of the model.

ASV: (Estimate=0.01388, p=0.052) appears to be marginally statistically significant but does not result in a negative change in the log INR. This is also illustrated in the graph in figure 4(c) where the decline in INR observed with different doses of ASV is like the decline over time, indicating that it provides no measurable benefit compared to time. Based on this, there is no clear benefit of administration of ASV at any dose. Considering the risk of anaphylaxis, harm clearly outweighs any benefit. This finding supports the guideline against administration of ASV in cases of confirmed envenoming by Hump-nosed pit viper.

Blood Products: Administration of blood products appears to have the largest and statistically significant change in log INR values (Estimate = 0.55368, p=<0.001). This finding is also corroborated by the chart. While blood products have been administered for the patients with the highest INR values, the transient vertical jumps in INR lines suggest that factor replacement temporarily results in lowering INR, but without clearance of venom, the INR increases. This finding also supports the guideline recommendations of using blood products cautiously, as a temporizing measure in case of bleeding manifestations. While blood products are an important component of supportive care, they only serve as a bridge to natural clearance of venom over time.

Therapeutic Plasma Exchange (TPE): (Estimate = 0.087, p = 0.656) As per the model, the effect of TPE is unclear. While the chart shows that TPE was initiated only for cases with very high INR values, it does not show a clear benefit. The decline in INR follows a similar trend to decline over time. However, this result must be interpreted with caution due to the very small sample size (of cases that underwent TPE), confounded additionally by delayed initiation and administration of blood products prior to TPE. Theoretically, while replacing clotting factors, TPE also removes circulating venom proteins. For severe envenoming in confirmed HNPV envenoming early TPE is a high-risk intervention that may save the life of a patient before they develop life threatening or permanent disability due to known complications such as stroke and acute kidney injury. Clinically as we observe a resolution of VICC with timely initiation of TPE, our center continues to use TPE in managing cases with severe envenomation (Fig 5). Only a few centers in the region have the resources and expertise to carry out TPE.

Considering the cost, resource, expertise requirement and morbidity associated with TPE, this study highlights the urgent need for an effective monovalent or regional polyvalent antivenom that is effective against HNPV envenomation.

### Diagnostic kits and antivenoms of the future

In current practice diagnosis is based on the availability of multiple evidence systems and often is only circumstantial. There are currently no POC diagnostic kits to determine species responsible for envenoming. Furthermore, current literature does not indicate any inclusion of HNPV in diagnostic kit development which is the need of the hour. It remains extremely challenging to differentiate envenoming between Russell’s viper, Hump nosed pit viper, saw scaled viper and Malabar pit viper without evidence of the biting species. This leads to longer therapeutic times as standard practice mandates the use of ASV which will not work in HNPV envenoming. In critical cases this delays the decision time to TPE and/or dialysis in reducing the morbidity and mortality of HNPV envenoming.

### Conservation and One Health approach

Hump-nosed pit vipers occur widely across their reported range in this region, from coastal forests to the hills of the Western Ghats. Being a forest species, it is often found on the forest floor, often camouflaged in leaf litter, occasionally seen climbing vegetation. They are currently facing pressure from human activity and land use patterns, with larger tracts of forests being modified for human use. This is likely to result in increased conflict in the region.

Adopting a one health perspective that integrated human health, wildlife conservation and ecosystem sustainability, our team coordinated with the Forest department to ensure the safe relocation of live specimens brought to the hospital. During the study period, a total of five live hump-nosed pit vipers were brought to the hospital by patients or attendants and were subsequently released back into their appropriate habitat close to where they were found following appropriate documentation and expert verification. This collaborative approach reinforces conservation-sensitive practices in snakebite management.

## Limitations

While this study adds valuable information on HNPV envenomation, it has many limitations including the small sample size, which results in low power of the linear mixed model analysis. As there is a lack of standard guidelines, there was no standardization in treatment practices.

## Conclusions/Recommendations

- This study confirms to emergency healthcare teams, wilderness experts and researchers about the burden and medical significance of Hump-nosed pit viper envenoming in this region and proposes an approach to its clinical management. (Fig 7)
- Specific or polyvalent ASV efficacious against HNPV venom in India is the need of the hour.
- TPE can be attempted in severe HNPV envenoming, however further study is required to assess the risk versus benefit of it.
- Areas with high risk of severe HNPV envenoming must have dedicated teams invested in awareness, education and research including venomics, anti-venomics and genetic profiling of the HNPV.

**Fig 7:**
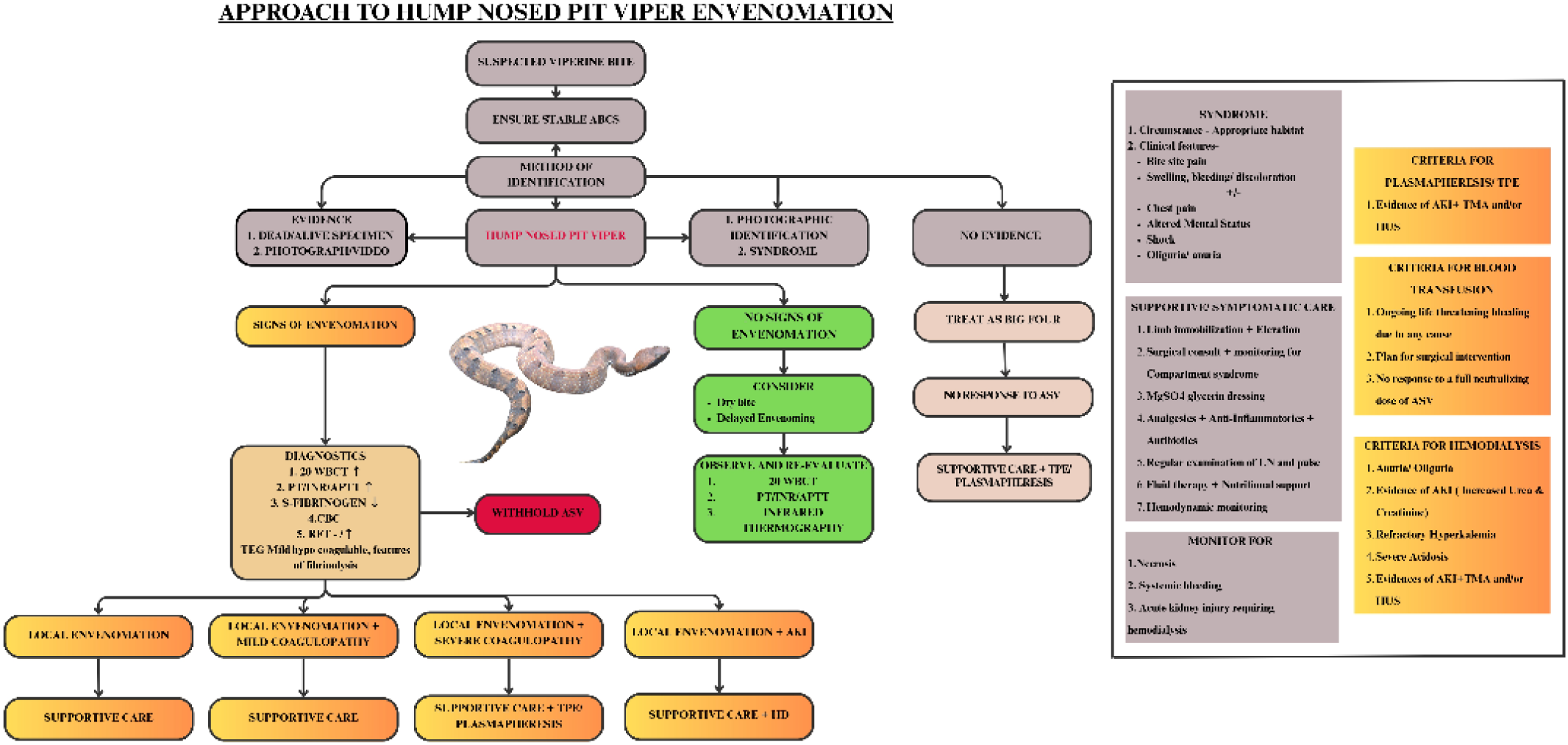
Approach to clinical management of Hump nosed pit viper envenomation.

## Data Availability

The data will be made available upon request

## Acknowledgments

We would like to thank Department of Emergency Medicine, Centre for Wilderness Medicine, KMC Manipal, Department of Emergency Medical Technology, Manipal College of Health Professions, MAHE, Manipal, Centre for One Health - National Centre for Disease Control, Herpactive and Mr. Alex Carpenter and for their constant support.

## Supporting information Captions

S1 Dataset: Raw dataset used for analysis

